# Responsiveness of functional assessments to monitor change in balance, walking speed and strength of older adults: A systematic review of the minimal detectable change

**DOI:** 10.1101/2022.06.06.22276029

**Authors:** Marco Arkesteijn, Daniel Low

**Author notes:** (Corresponding Author) Dr Marco Arkesteijn.

## Abstract

**Objectives:** The objective of this paper is to systematically review and evaluate the responsiveness of different functional tests via the minimal detectable change (MDC) across different older adult population cohorts.

**Design:** Systematic review of ISI Web of Knowledge and PubMed databases were searched up to September 26th 2020.

**Setting:** Community dwellings, hospital and residential homes

**Participants:** Studies were included if participants were adults over the age of 60. This study reports data from studies that utilise healthy community dwelling older adults, as well as older adults who are hospitalised, live in residential home or have musculoskeletal conditions.

**Interventions:** No interventions feature in this study

**Primary and secondary outcome measures:** MDC reported for gait speed, grip strength, balance, timed up and go, and repeated chair stand separated per older adult sub-group were deemed the primary outcome measure. A secondary outcome measure were the results of a regression analysis, performed to determine the effect of the functional test, cohort, study design and MDC calculation methodology on MDC magnitude.

**Results:** Thirty-nine studies met the inclusion criteria. Not all assessments were evaluated in the literature for all population cohorts. The MDC was affected by the functional test used, the cohort and MDC calculation methodology.

**Conclusion:** The MDC can be assessment and population specific, and thus this should be taken into account when using the MDC. It appears acceptable that different assessors are involved in the re-assessment of the same person.

**Trial registration:** The systematic review protocol was published in PROSPERO (CRD42019147527).

**Article Summary:** *Strengths and Limitations of this Study:*

- Strength: A range of assessments were included to determine if MDC could be used to prioritize specific assessments in interventions.
- Strength: A wide range of search criteria and methods resulting in 6448 studies being assessed that enabled the inclusion of 39 original research papers to derive 138 MDC values.
- Strength: Analysis of MDC_95_ for functional tests commonly used by practitioners to assess effective change in older adults
- Strength: Analyses of the impact of method design features such as different assessors on the MDC
- Limitation: Limited to the settings and tests selected

## Introduction

The aging process can result in various physiological and biomechanical changes ^1-3^, which can lead to, or result from the onset of frailty ^4^, or diseases such as osteoarthritis ^5, 6^ or sarcopenia ^1, 3^. In particular, co-morbidity such as musculoskeletal conditions and acute and chronic living conditions such as hospitalisation and residential home habituation respectively can exacerbate the change that occur due to aging^7^. The changes underlying physiological, biomechanical and cognitive processes can negatively impact functional ability of the individual. Recently, the World Health Organisation defined functional ability as the combination of the intrinsic capacity of the individual, the environment a person lives in and how people interact with their environment^8^. This includes walking, balance and strength as indicators of intrinsic capacity^8^. Subsequently, a decrease in the ability to perform these activities will affect their quality of life and well-being^4^. To monitor an individual’s function ability, functional performance assessments are used to establish change in functional ability due to aging, that predict increasing mortality risk, falls likeliness, and cognitive decline ^2, 9-11^.

Changes in functional performance can reflect variances in cognitive processes ^2^, neuromuscular control ^10^, endurance ^12^, muscle strength and ‘physical power’ ^10, 13^, and poorer static and dynamic balance ^14^. These changes can lead to fear of falling ^15^, perception of social isolation/participation ^16^, an inability to perform daily activities and immobility ^17^ and a generally reduced quality of life ^16^. Therefore, it appears important to monitor changes in functional ability over time, both across the lifespan due to aging and during acute events. Similarly, it is important to monitor those changes occurring between the short and long term, such as those due to hospitalisation, musculoskeletal conditions or residential home living. This allows monitoring of the deterioration in patient condition and the responsiveness to interventions by clinicians and therapists.

Various functional performance assessments exist that are safe, cost-effective, require limited time and equipment, and are easy to interpret ^18^; these tests include the five-times or 30-second Sit to Stand (5xSTS and 30sSTS respectively), Hand Grip Strength (HGS), Berg Balance Scale (BBS), Six Minute Walk (6MWT) and Two Minute Walk (2MWT), normal (NGS) and fast gait speed (FGS), Timed Up and Go (TUG), and Single Legged Stance (SLS). These tests are commonly used to measure functional capacity in both clinical and community-based settings for a range of populations ^19^. In addition, their scoring is primarily based on continuous data, meaning that minor changes might be identified. This is in contrast to assessments involving cut offs to score performance, such as the Short Physical Performance Battery (SPPB). Despite being popular, the SPPB has the inherent limitation that for example an increase in chair stand test duration of 2.5 seconds might not lead to a change in performance if the baseline was 14.0 seconds. Thus, particular functional assessments employing continuous data for scoring can provide valuable insight into the monitoring of patient centred outcome measures while being objectively quantified.

The ability to observe change or difference in functional assessments is influenced by the size of error within observed values ^20^. This error can be indicated by the absolute reliability of the data, demonstrated via the calculation of the Minimal Detectable Change (MDC) ^21^. This statistic provides a quantifiable value that defines the responsiveness of the functional assessment; a lower MDC suggests a better ability to detect a small improvement or deterioration in functional ability ^21^.

Published systematic reviews exist, which detail the responsiveness of gait speed tests in older adults ^22-24^. These reviews do not however consider potential differences between normal gait speed and fast gait speed. Similarly, Downs et al. (2013) ^25^ reviewed the literature for the BBS test, reporting a curvilinear relationship between the average BBS score and the MDC statistic. The MDC of 2MWT ^26^, HGS ^27^, and 6MWT ^28^ have been shown to vary across patient groups suggesting that the responsiveness is population/health condition specific. It is possible that the inter-session reliability of assessments is impacted by the health condition of the patient, such as daily variations in health, pain and subjective feelings ^29^, whereas this is less likely to affect the intra-session reliability. Thus, musculoskeletal conditions as well as the impact of hospitalisation and residential care living could increase task performance variability compared with healthy individuals. This would increase the MDC, and mean that condition specific MDC values should be used over MDC values calculated on other clinical populations or from the general population. This is a factor not considered in the interpretation of the MDC within the aforementioned gait speed and BBS reviews. Many of these reviews also analyse the data for all age groups without exploring the MDC specifically for older adults. This population’s physiological changes may add further noise to that present due to a specific health condition and increase the MDC reported. These reviews have also excluded studies with small sample size (n < 10) ^25^, which along with methodological considerations such as the Standard Error of the Measurement (SEM) calculation method used in the MDC calculation, will impact the data reliability statistics ^30^. In addition, the length of time between trials will also likely impact data noise and the MDC calculated and these factors need to be understood when exploring a suitable MDC to be used when evaluating change.

The purpose of this review was to systematically search the literature to provide an updated review of MDC statistics for the 5xSTS, 30sSTS, BBS, 6MWT, 2MWT, TUG, SLS, HGS and NGS and FGS for healthy older adult populations as well as populations who can be defined as having musculoskeletal conditions, hospitalised or in residential home. This will enable the generation of recommendation for clinicians and therapists to monitor patients over time, and for example determine the individual effectiveness of rehabilitation programmes. Secondary aims were to determine the impact of the study design, SEM calculation method, time between trials or assessor scoring and population size on the MDC measurement. In addition, since the selected functional tests measure similar physiological characteristics, the impact of the test chosen on MDC_95_ was also explored.

## Methods

### Data Sources and Searches

The systematic review was designed collectively by two researchers. ISI Web of Knowledge and PubMed databases were searched using the terms identified in Table 1 for all available dates up to September 26th 2020. Subsequently, manual searches from relevant systematic reviews included in the database search and references lists of included manuscripts were performed. This systematic review protocol was registered with PROSPERO (Registration Number CRD42019147527).

**Table 1.** Participant, Test and Outcome search terms used for systematic review

| Terms used |
| --- |
| Elderly OR aging OR ageing OR old* OR geriatric |
| Minimal Detectable Change OR MDC OR SEM OR Standard Error<br>OR Reliability OR ICC OR intraclass correlation coefficient OR<br>Minimal important change OR Minimally important change OR MIC<br>OR Smallest Detectable Change OR SDC OR Smallest Worthwhile<br>Change Or SWC |
| Sit to Stand OR chair-stand OR chair-rise OR Berg Balance Scale OR<br>Balance Scale OR Six Minute Walk OR Minute Walk OR Timed Up<br>and Go OR TUG OR Single-leg stance OR Single-legged stance OR<br>gait speed OR sit and reach OR Chair sit and reach OR flexibility |

### Study Selection

Eligible studies were those which reported data for study participants with a mean age greater than 60 years. Eligible studies also included those which reported the SEM or the MDC or analogous terms (Smallest Real Difference (SRD) or Smallest Detectable Change (SDC)). These were identified for the following functional assessment measures: 5xSTS and 30sSTS, BBS, HGS, 6MWT, 2MWT, TUG, SLS, FGS and NGS. Intervention studies, defined as a study which implemented a strategy to change outcome measures over a period of 3 or more weeks, were included only if the MDC and/or SEM data was established using test and retest data collected pre-intervention or post intervention. Studies were also included if it showed an inter-rater (at least two assessors rating a patient’s single performance at the same time on 1 occasion), intra-patient/inter-rater (at least two assessors scoring the patient’s performance on different occasions), and patient test-retest (a patient’s performance was assessed on different occasions by the same assessor) design. Those studies which did not detail the study design or which the design could not be classified as one of the above were excluded, as were those which were not original research studies (i.e. used the data published elsewhere) or were not written in English. From the extracted data, the present study focussed on studies comprising of healthy community dwelling, musculoskeletal conditions, hospitalisation or residential care home participants.

For the papers retrieved, the two authors of this paper independently screened the titles and abstracts for the inclusion criteria. Any research that was not clear whether the criteria was met, underwent a review of the full text and was accepted or rejected accordingly. The reference lists of the accepted studies also underwent the same review process and any additional research publication which met the search criteria and which were not found from the initial database search were then included. Finally, a check of similar systematic reviews also helped to ensure relevant sources were not missed.

### Data Extraction and Quality Assessment

Information on the patients’ health status (healthy community dwelling, musculoskeletal conditions, hospitalisation or residential care home), the number of patients sampled and the statistical approach used to calculate the SEM and/or MDC value were extracted into a central data sheet. Additionally, the mean or median average for each cohort on the first trials (baseline) were extracted (or group mean if these values are not provided), along with time between trials or assessors scoring. When required, data were converted to ensure the same measurement units in MDC value across studies (i.e. cm/s to m/s). Likewise, to ensure comparable statistics, when MDC were reported using the z-score associated with a 90% Confidence Interval, these were converted to 95% using the SEM reported in their study or by using Equation 1. All data entered into the central sheet were checked for accuracy by both authors.

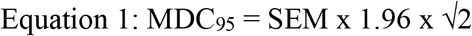

### Data Synthesis and Analysis

The MDC_95_ values were averaged across all studies for each assessment and their range was determined. Extracted MDC_95_ values were also transformed into a percentage of the baseline assessment score if this value was not provided by the study, to enable comparison of the magnitude of the MDC_95_ across assessments (MDC_95_%). In line with Chapter 10 of the Cochrane Handbook for Systematic Reviews for Interventions (http://www.training.cochrane.org/handbook), meta-regressions can be used to investigate differences in an outcome variable changes for continuous and categorical explanatory variables. Therefore, to explore the variation in the MDC_95_ data, an Enter Method multilinear regression was performed using the MDC_95_% as the dependent variable.

Functional assessment, SEM calculation method (use of a one-way random, two-way random or two-way fixed ICC or square root of mean square error (√MSE)), study population, number of study participants, time between trials or assessor scoring (1 day or fewer, within three days, within seven days, seven days specifically, within two weeks) and study design (intra-rater/intra-patient, inter-rater/intra-patient or inter-rater) data were then used as the independent variables. For time between trials or assessor scoring and the SEM calculation method variables, any category with less than five studies were combined with those which provide unclear data and grouped as ‘other’; these were included in the analysis since these studies include information relating to the other independent variables. However, these ‘other’ groupings were excluded from later interpretation. Since the functional assessment, SEM calculation method, study population, time between trials or assessor scoring and study design data were categorical, the different categories were entered into SPSS as separate columns. Dummy variable (i.e. 1 = yes or 0 = no) were then used to indicate the presence of the independent variable category (level) in the calculation of the MDC_95_ statistic. To obtain beta coefficients for each level, multiple regression analyses were performed, with each level within the independent variable used as a reference variable in one of the analyses (e.g. not placed into the regression).

Beta coefficients are reported for all variables, along with the r and r^2^ value and F-value from the ANOVA table. Significant contribution of the independent variables to the variation in dependent variable was indicated at the 95% confidence level when p < 0.05. Comparisons between groups were expressed as absolute difference in MDC_95_%, thus in percentage points difference in MDC_95_%.

## Results

A total of 39 studies reporting the MDC_95_ or provided SEM data to calculate MDC_95_ were included in this review (Figure 1). Table 2 shows the frequency of MDC_95_ for each assessment per population. Out of the 39 studies reporting the MDC_95_, a healthy, community dwelling older adults was available for all assessments except for the 30sSTS, while the MDC_95_ was less frequently available for specific populations for other assessments. The average and range of MDC_95_ values for each assessment are shown in Table 3 and Figure 2. A full description of the included studies is provided in Appendix A.

**Figure 1.**
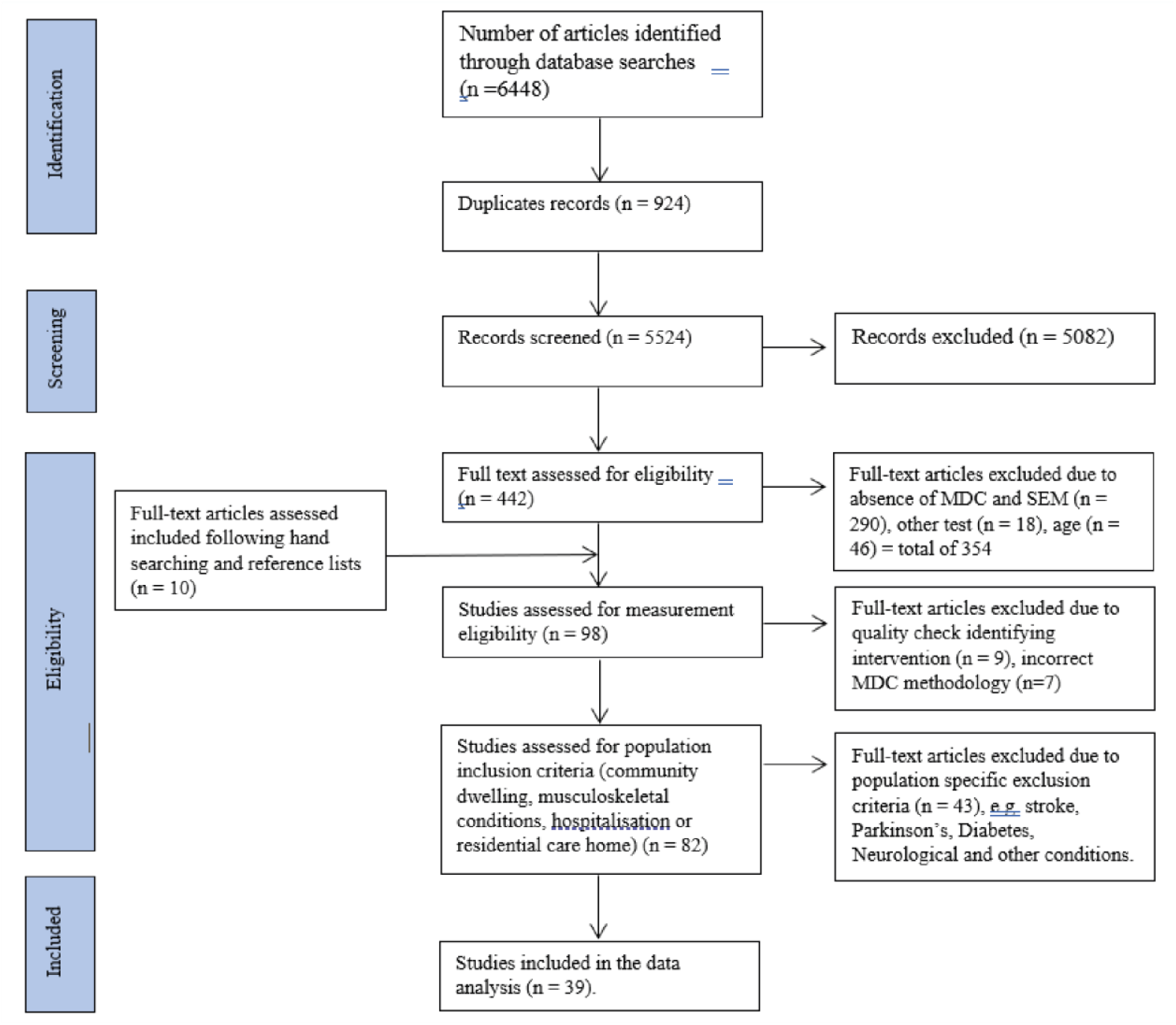
Flowchart of the systematic review and meta-analysis according to the PRISMA guidelines.

**Table 2.** Summary of the availability of population specific MDC95 value for the five-times and 30-second Sit to Stand (STS5x and STS30s respectively), Timed Up and Go (TUG), Hand Grip Strength (HGS), normal (NGS) and fast gait speed (FGS), Six Minute Walk (6MWT) and Two Minute Walk (2MWT), Single Legged Stance (SLS) and Berg Balance Scale (BBS).

| Assessment | Number of studies | Healthy, community-dwelling | Hospital | Residential | Musculo-skeletal |
| --- | --- | --- | --- | --- | --- |
| STS30s | 12 | 1<sup>38</sup> | 1<sup>39</sup> | - | 2<sup>40, 41</sup> |
| STS5x | 13 | 3<sup>42, 43, 44</sup> | 1<sup>45</sup> | 1<sup>46</sup> | 3<sup>47, 48, 49</sup> |
| TUG | 37 | 7<sup>50, 51, 52, 53, 44, 54, 55</sup> | 1<sup>54</sup> | 1<sup>46</sup> | 8<sup>56, 57, 41, 58, 59, 60, 47, 61</sup> |
| SLS | 5 | 2<sup>42, 62</sup> | - | - | 1<sup>31</sup> |
| GS | 13 | 4<sup>63, 42, 64, 55</sup> | 1<sup>39</sup> | 1<sup>46</sup> | 2<sup>63, 65</sup> |
| BBS | 11 | 1<sup>51</sup> | - | 1<sup>66</sup> | - |
| NWS | 19 | 5<sup>42, 67, 50, 46, 68</sup> | 2<sup>69, 70</sup> | 1<sup>46</sup> | 2<sup>71, 65</sup> |
| FWS | 5 | 2<sup>50, 72</sup> | - | - | - |
| Six mw | 22 | 1<sup>50</sup> | - | - | 4<sup>57, 73, 41, 60</sup> |
| Two mw | 11 | 2<sup>42, 74</sup> | - | 2<sup>75, 74</sup> | 1<sup>59</sup> |

**Table 3.**
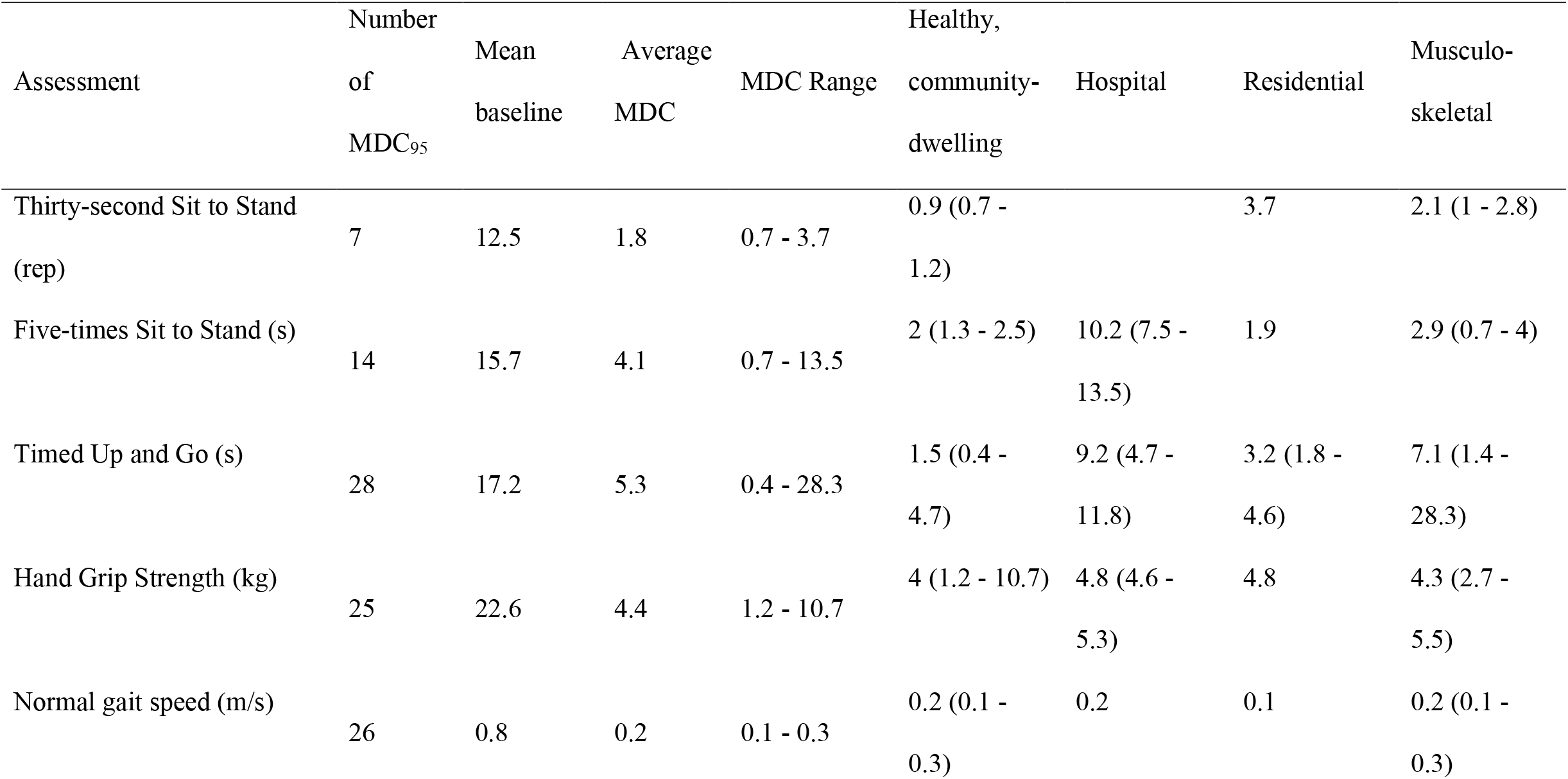
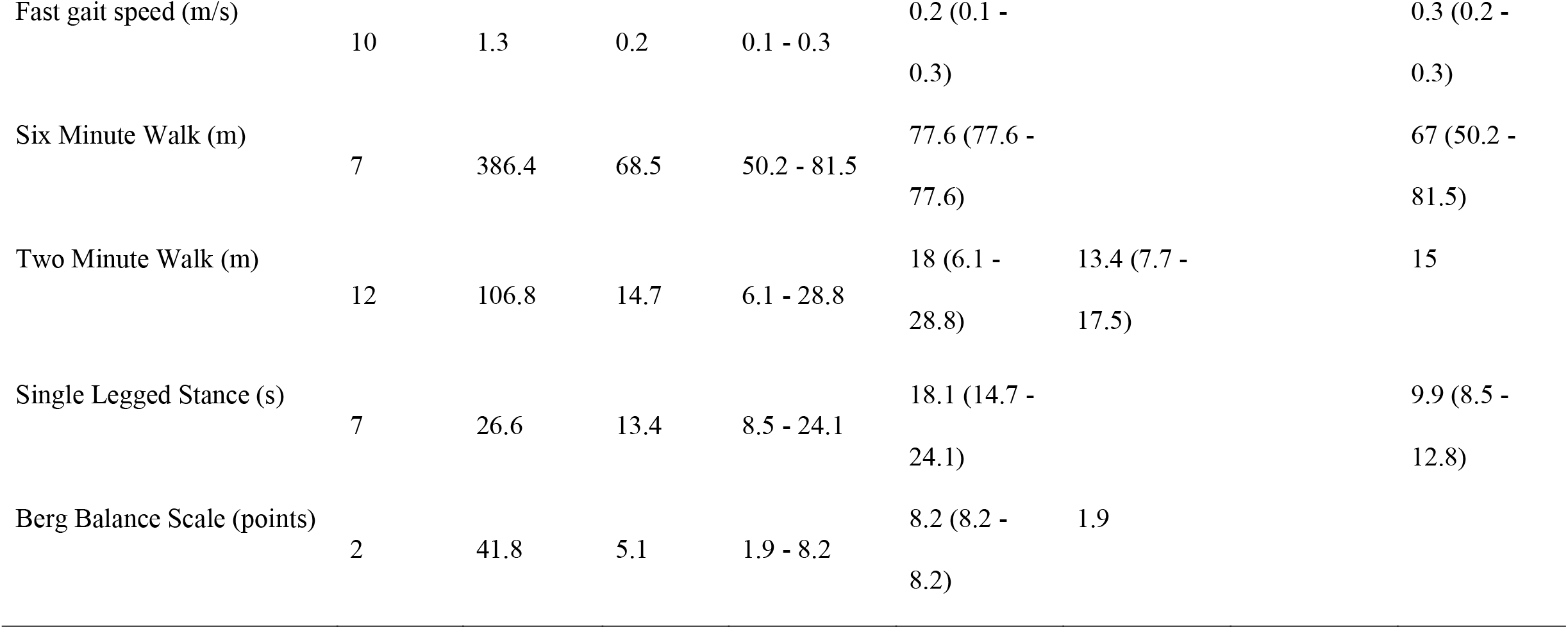
Summary of the average Minimum Detectable Change (MDC_95_) and range for each functional assessment in absolute values. Empty cell indicate no MDC_95_ was present. Populations specific values are average, and in brackets the range if multiple values were present.

**Figure 2.**
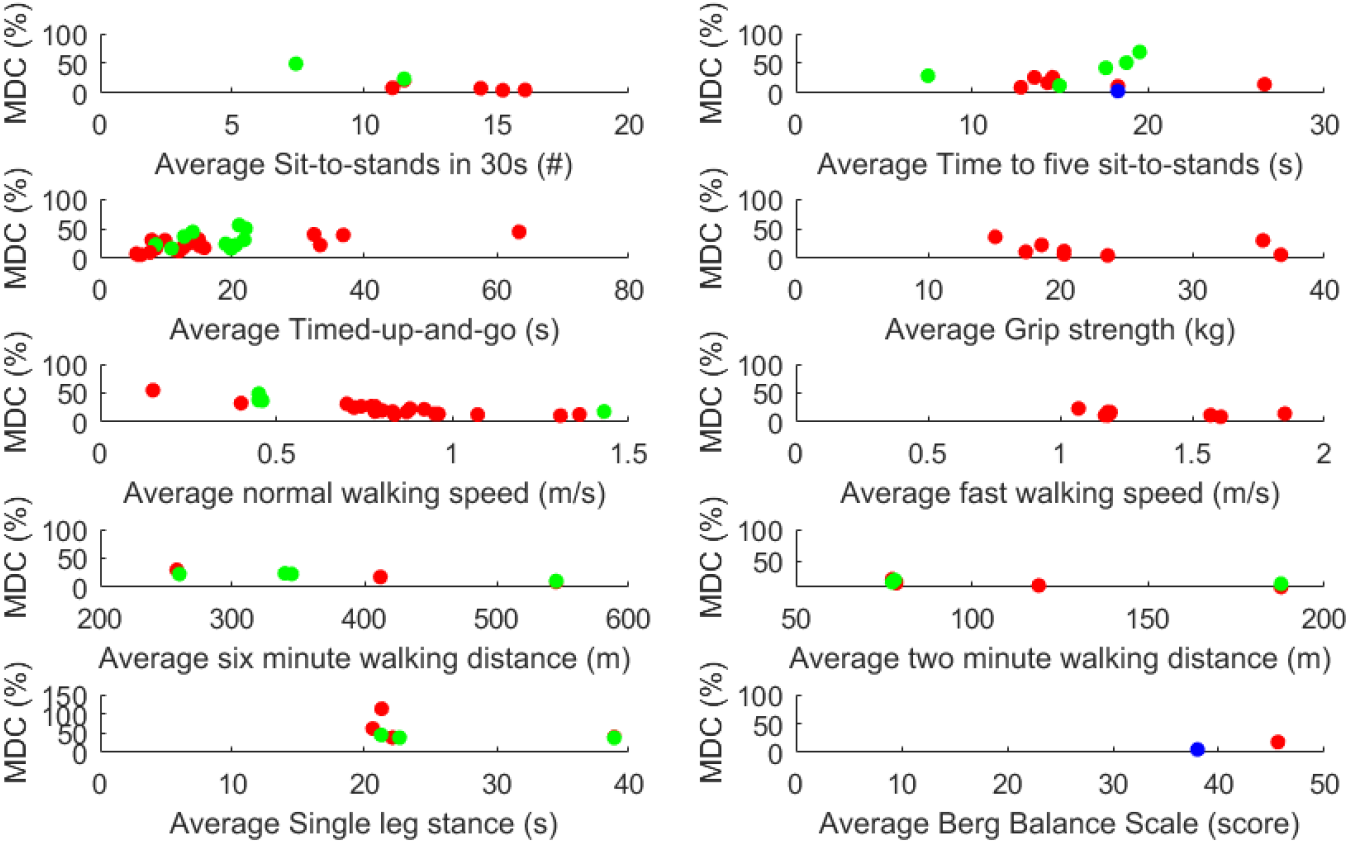
The distribution of Minimum Detectable Change (MDC) values for each assessment as a function of the average score. Red colour indicates MDC values from patient test-retest design, green colour indicates an intra-patient/inter-rater design, and blue colour indicates an inter-rater design. Note: The vertical axis of the Single Leg Stance comprises 0-150%, rather than 0-100% range observed in other assessments.

There were 138 MDC values included in the regression analysis. The following number of values were therefore included in the regression analysis (note, some studies reported more than one MDC_95_ value): Functional test (FGS (n = 10), NGS (n = 26), TUG (n = 28), HGS (n = 25), 5xSTS (n = 14), 30sSTS (n = 7), SLS (n = 7), BBS (n = 2), 6MWT (n = 7), 2MWT (n = 12); Design (intra-rater/intra-patient (n = 97), inter-rater/intra-patient (n = 39); inter-rater (2)); SEM calculation method (one-way random effects (n = 2), two-way random effects (n = 27), two-way mixed effects (n = 7), mean square error (√MSE) (n = 51, other/unknown (n = 51)); Time between trials or assessor scoring (1 day or fewer (n = 44), within 3 days (n = 20), within 7 days (n = 20), 7 days (n = 21), within 2 weeks (n = 21), unknown/other (n = 12)); Population (residential (n = 25), musculoskeletal (n = 63), hospitalised (n = 9), healthy community dwellers (n = 41)).

The results of the regression analysis indicated a significant correlation (R = 0.74, p < 0.01) with the r^2^ indicating that model explained 55.1% of the variance in the dependent variable and was a significant predictor of MDC_95_%, (F(24,113) = 5.777, p < 0.001). Variation in the MDC_95_% could be explained by the functional test and sample population used, and the time between trials or assessor scoring (p ≤ 0.05). Conversely, the SEM calculation method and the study design (patient test-retest or intra-patient/inter-rater) was not a significant predictor of MDC_95_% magnitudes although inter-rater studies were smaller than intra-patient/inter-rater (21.085MDC_95_% points). Similarly, the regression analyses demonstrated that there was no significant effect of the number of patients sampled on MDC_95_% (beta = -0.013 years, t = -1.133, p = 0.260). A full description of beta-coefficients are provided in Tables 4-8 (Appendix A).

The analysis demonstrated that the population used to calculate MDC_95_% impacts the size of the MDC_95_%. More specifically, those who were healthy community dwellers had smaller MDC_95_% values compared to residential patients (10.728MDC_95_% points). The regression also showed differences between the functional test being used, whereby all functional tests possessed significantly smaller MDC_95_% than the SLS (30sSTS (39.440MDC_95_% points), HGS (37.941MDC_95_% points), 5xSTS (32.386MDC_95_% points), BBS (36.156MDC_95_% points), NGS (33.221MDC_95_% points), FGS (39.460MDC_95_% points), 6MWT (38.207MDC_95_% points), 2MWT (44.240MDC_95_% points) and TUG (30.842MDC_95_% points)). In addition, the 2MWT MDC_95_% was smaller than that of the TUG (13.398MDC_95_% points) and the HGS MDC_95_% was smaller than the NGS test (4.720MDC_95_% points). Further still, when the data were collected within a day of each other, the MDC_95_% was smaller than when collected within a week of each other (12.139MDC_95_% points); those collected within seven days were also larger than those collected specifically in 7 days (13.098MDC_95_% points)

## Discussion

The present systematic review has identified that there is a large range of MDC_95_ values reported across studies. It is the first study to confirm factors that influence the MDC_95_ include the cohort population and the type of functional test being performed. In addition, aspects of the study procedures, such as the time between trials and the study design also contribute to variation in the MDC_95_. These results provide additional support for specific cohort population requirements when choosing the MDC_95_, which allows clinicians and therapists a more considered choice of test to use with their clients.

The present systematic review is the first to determine that the MDC_95_ differs between functional tests. For balance assessments, the SLS possessed a greater MDC_95%_ magnitude than BBS and all other assessments. Consequently, this highlights that the SLS is not recommended for use to monitor individuals, although this disagrees with Choi et al. ^31^. A potential reason could be that Choi et al. ^31^ based their interpretation on data utilising higher ICCs and was specific to osteoarthritis patients.

Similarly, for gait speed assessments, the TUG and 2MWT were different, whereby the 2MWT possesses a smaller MDC_95_%; the values for TUG, 6MWT, FGS and NGS which can also be used to measure walking speed, were however similar. It is important to clarify that tests such as the TUG, 2MWT and 6MWT are indicative of endurance and mobility that would impact walking speed, whilst FWS and NWS are more direct measure of typical walking speed. These findings may suggest that the 2MWT introduces less error than TUG. Alternatively, it may suggest less sensitivity to calculation errors when the 2MWT is used. The MDC_95_% of FGS was also not significantly different to that of NGS, a comparison not previously explored ^22-24^. These results therefore highlight the importance of considering the test being used to assess patients. It suggests the 2MWT may offer the greater ability to detect smaller changes than the other measures, although it is acknowledged that this will be quite time-consuming and requires substantial space requirements. In contrast to balance and gait speed assessment, lower leg strength as indicated via the tests TUG, 30xSTS and 5xSTS provided similar MDC_95_% indicating equal ability to detect differences between population groups.

The present systematic review also confirms that there is variation in the MDC_95_ across patient populations, as suggested previously for 6MWT ^28^, 2MWT ^26^ and HGS ^27^. The multivariate regression showed that fluctuations in daily performance appear greater in individuals with underlying health conditions compared with healthy individuals. These relative differences will likely be due to the different levels of noise within the data, a result of the differing health conditions and its effect on the neuromuscular functioning of the body ^32^. However, despite the increased number of studies reporting the MDC compared to previous systematic reviews, Table 2 shows that not all populations selected in this study have been evaluated to date for multiple functional tests. Therefore, clinicians and therapists need to consider whether further reliability study is required for these population-functional test combinations. Similarly, under-powered comparisons may prevent other differences between populations from being demonstrated in this study.

The highest MDC_95_% values were found to be within 7 days between assessments, with generally around 10 MDC_95_% points higher than assessments done on the same day. It remains unknown why MDC_95_% is increased between 1-7 days, but speculatively might be due to regular activities varying from day to day, but repeated weekly thus leading to similar values when at a 7-day separation period was used, which was also different to data collected within a week of each other. MDC_95_% around 1 week offers some support to the view of Choi et al. (2014) ^31^, who recommended that 7 days between trials as this was long enough for a learning effect to be mitigated but short enough for clinical status not to change. Yet, since there was no difference in MDC_95_% between those collected on the same day with those with those with at least 7 days separation, it would seem advantageous to collect multiple trials on the same day.

The regression analysis further revealed that there was no effect of repeated measure study design, thus having the same assessor to measure the test and retest appears to have the same impact on the MDC_95_ as the use of different assessors. This in agreement with some individual studies (e.g. ^33, 34^), but potential limitations of this study could be because of publication bias and large heterogeneity of study methodologies included in the present review. The lack of effect of assessor was likely related to the simple nature of the tests, sufficient assessor training and the use of the same or similar equipment. Consequently, the MDC_95_ in the studies identified mostly reflects error due to performer. Combined, it appears that future intervention studies should consider having multiple assessments on the same day or between 7 days apart, but this does not necessarily have to be the same assessor.

The results of the regression analyses also indicate that the ICC model had little impact on the MDC_95_ and did not find evidence to suggest that the two-way fixed model data should not be used beyond the study in which it has been conducted ^30^. Compared with the two-way fixed model, the two-way random model uses the systematic and random errors as separate sources of error in the ICC calculation. Consequently, given the presence of systematic error which is of similar magnitudes, the two-way fixed model will underestimate the MDC_95_ calculated ^30^. However, given the similar magnitudes it may suggest that the level of systematic error is low and thus not impacting the values obtained. Further still, the √MSE calculation did not result in a lower MDC_95_ compared to the two-way random model or two-way fixed model. In comparison to the two-way random model, this was unexpected, whereas in comparison to the two-way fixed model this was as anticipated given that systematic error was also ignored by this method of SEM calculation. The one-way model produced similar MDC_95_% to those calculated by two-way models. These observations are unexpected since the one-way model should offer a more conservative reliability statistic and thus a larger MDC_95_ when errors are similar ^30^. A potential explanation could be that low statistical power and varying magnitudes of random and systematic errors across studies, due to different method considerations, may have had a larger impact on the difference in MDC_95_% between these models, especially since the number of data sets included for the one-way model was relatively low (n = 16).

From a practical implementation perspective, clinicians and therapists should consider the population and MDC calculation techniques before using the currently available MDC values and that differently designed analyses might not provide appropriate MDC values. Moreover, there was a large range of MDC values across studies and the regression model only explained 54.4%, suggesting that many other factors could limit the use of pre-existing MDC values. Further research to better understand variability of assessments is thus warranted. In addition, the typical MDC_95_ exceeding 20% suggests that monitoring an individual’s change requires on average at least 20% change if done with a single assessment. This for example equals around 7 or 15 years of ‘typical’ decline for hand grip strength for an 80 or 60-year-old person respectively ^35^, indicating that the MDC is considerably large. Therefore, future research could therefore also explore alternative techniques to the MDC to monitor the impact of the aging process on functional ability. This could improve the ability to identify functional decline earlier, and intervene earlier in a person-centred approach, and thus potentially better long-term outcomes for the patient. A potential alternative approach would be to determine an MDC equivalent for each individual, based on that individual’s variation in repeated assessments. This would however require multiple measurements and would warrant ubiquitous monitoring, or self-assessment techniques to be developed that allow repeat measurements to detect automatically.

### Limitations

Limitations of this paper includes that no risk of bias assessment was performed, due to lack of existing tools ^36^. However, recently, a Delphi study made various recommendations ^36^, which are similar to the methodology employed in the present systematic review, confirming the quality of the data exclusion criteria. Nevertheless, in some studies, the method used were not clear meaning that for some independent variables they needed classification as ‘other’. The number of studies available in these populations may also suggest that the results for some independent variables led to underpowered comparisons and thus potential type 2 error in the conclusions. The study also reviewed data for specific cohorts of older adults, and given the effect of these on the results obtained, future research should summarise the impact of other health conditions on the MDC_95_ in older adult population; it would also be worthwhile to explore the impact of health conditions within younger population. Furthermore, the MDC only reflects the value needed for a difference to be beyond chance and not necessarily the magnitude of change needed for clinical symptoms to change ^21^. A recent expert panel on frailty and sarcopenia highlighted the importance of Minimal Clinical Important Change statistic to determine clinically meaningful change for physical performance measures ^37^, thus a systematic review of anchor-based values that indicate this would therefore be a useful further study. Finally, the present study did not focus on the minimal change required for groups, which can be quantified using the SEM and is effectively the MDC_95_ divided by 2.77 (see equation 1). Combined, a comparison of both the MDC and MCIC with effect size of intervention studies could increase the understanding of the magnitude and impact of an intervention.

## Conclusion

In conclusion, the MDC_95_ can be assessment specific and when using a previously established MDC one should consider the impact of the method used, particularly when one-way models or √MSE values are used in the calculation. Thus, clinicians and therapists should be careful about relying on a single MDC study for their interpretation of individual patient changes, regardless whether this is for ongoing monitoring or rehabilitation interventions. Gait speed measured via 2MWT appears to be less sensitive to, or result in less variation between trials than other measures of gait speed and the single leg stance is not recommended for use. It appears acceptable that different assessors are involved in the re-assessment of the same person. The minimal detectable change is dependent on the population of interest. However, for some assessments, population specific indicators of responsiveness are not currently available, and therefore the present systematic review provides a guide to appropriate values to employ as the minimal detectable change.

## Supporting information

Conflict of interest form

Supplemental file 1: Protocol details

Supplemental file 2: MDC Data

## Data Availability

All data is retrievable from the manuscript and supplementary files, including the search results via EndNote files.

https://osf.io/jc7b8/?view_only=a53734d68e20463c9b007efe60453711

## Acknowledgements

There are no acknowledgements for this study

## Author Contribution

Both authors were central to the conception, development of protocol, summary, analysis and discussion of data and the preparation of this manuscript.

## A data sharing statement

Endnote data file of all retrieved papers is shared via https://osf.io/jc7b8/?view_only=a53734d68e20463c9b007efe60453711

## Appendix A: beta coefficients

**Table 4:**
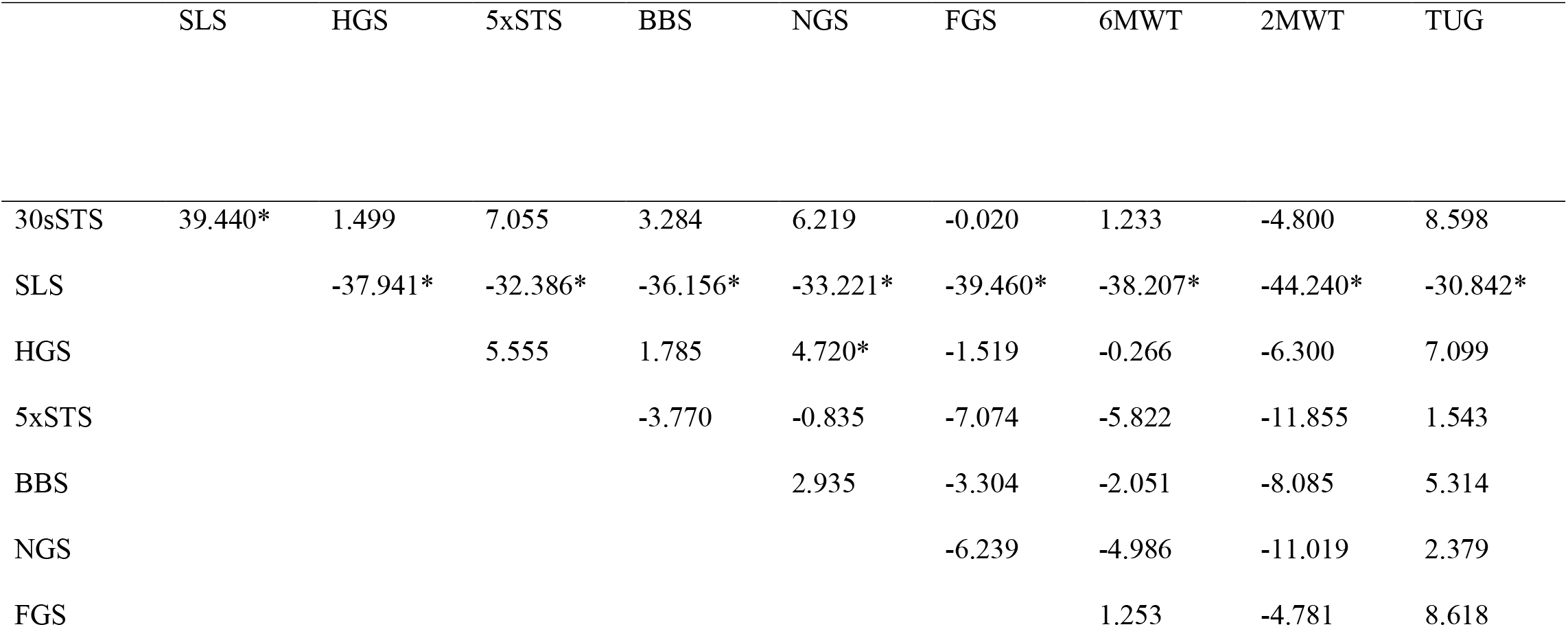
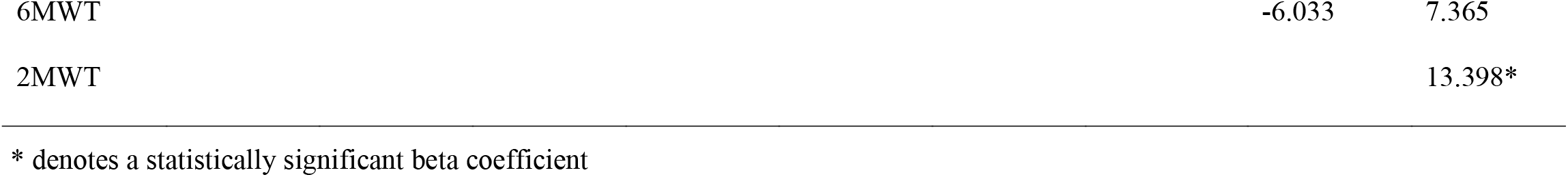
Multiple regression beta coefficients for MDC_95_% calculated for the 30 second sit to stand (30sSTS), single leg stand (SLS), handgrip strength (HGS), 5 times sit to stand (5xSTS), Berg Balance Scale (BBS), normal gait speed (NWS), fast gait speed (FWS), six minute walk test (6MWT), two minute walk test (2MWT) and Timed Up and Go (TUG). Vertically listed tests are the reference variables within the regression.

**Table 5:**
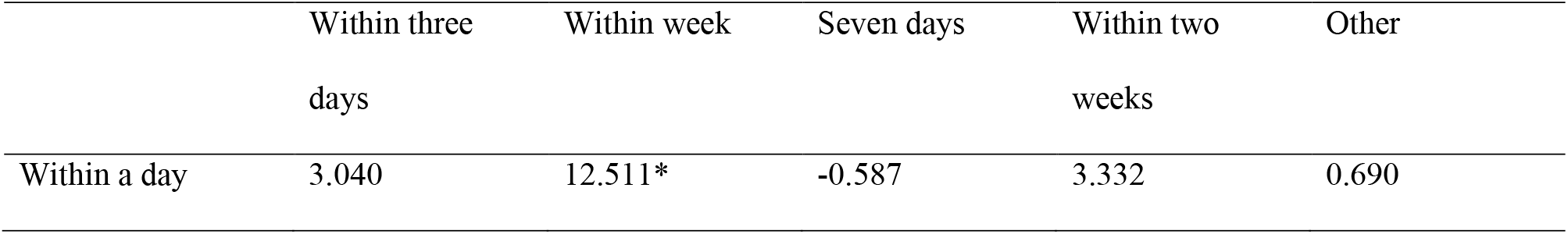
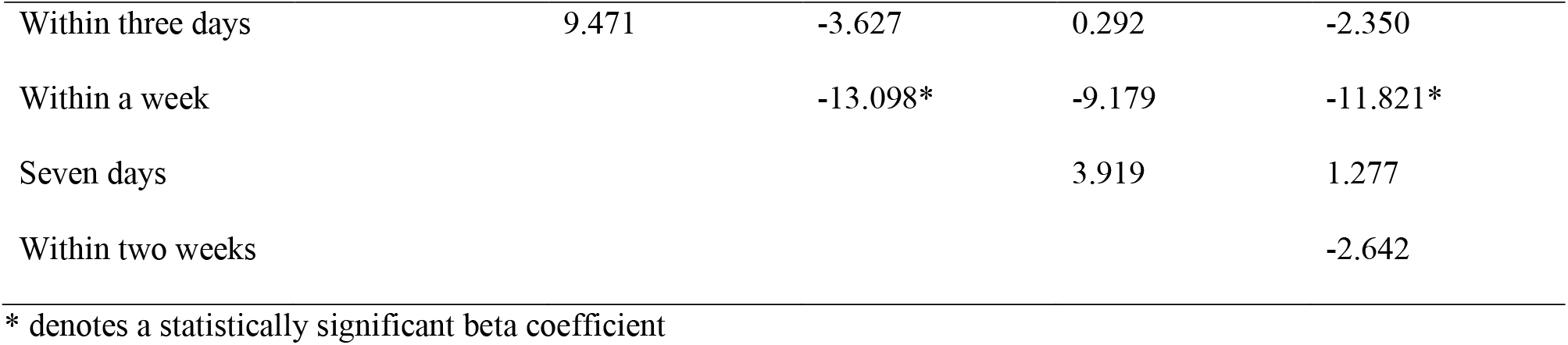
Multiple regression beta coefficients for MDC_95_% calculated with data collected within a day, within three days, within a week, seven days, within five to ten days, and within two weeks separation, as well as those collected with other time periods. Vertically listed time periods are the reference variables within the regression.

**Table 6:** Multiple regression beta coefficients for MDC_95_% calculated for studies with patient test-retest, intra patient/inter rater and other (inter-rater) designs. Vertically listed designs are the reference variables within the regression.

|  | Intra patient/inter rater | Inter-rater |
| --- | --- | --- |
| Patient test retest | 1.051 | -20.034 |
| Intra patient/inter rater |  | -21.085* |

\* denotes a statistically significant beta coefficient

**Table 7:**
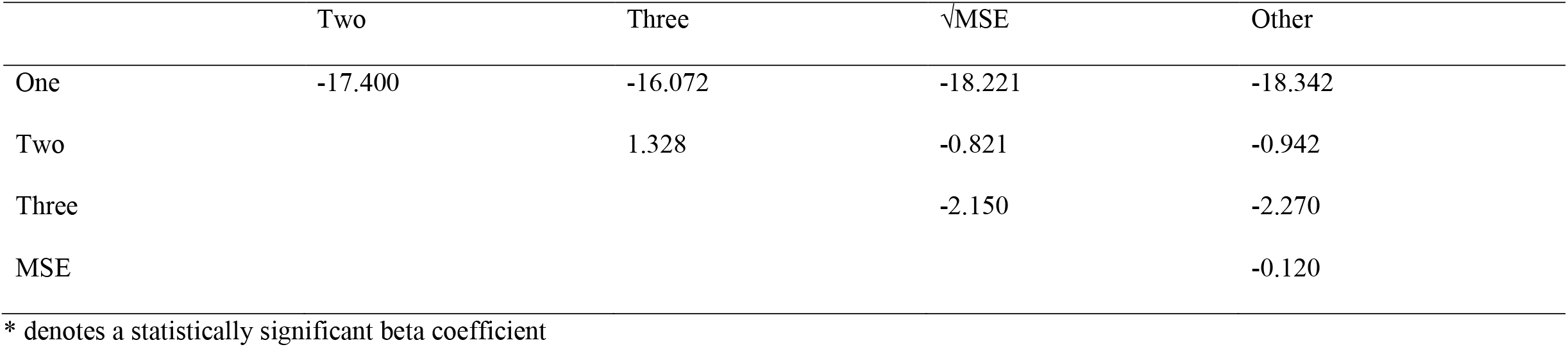
Multiple regression beta coefficients for MDC_95_% calculated using Standard Error of the Measurement determined with the ICC from a one-way random effects (One), two-way random effects (Two), or two-way mixed effects (Three) model or using the square root of mean square error (√MSE) or other approaches (Other). Vertically listed approaches used to calculate the SEM are the reference variables within the regression.

| | Two | Three | $\sqrt{\text{MSE}}$ | Other |
| --- | --- | --- | --- | --- |
| One | -17.400 | -16.072 | -18.221 | -18.342 |
| Two |  | 1.328 | -0.821 | -0.942 |
| Three |  |  | -2.150 | -2.270 |
| MSE |  |  |  | -0.120 |

\* denotes a statistically significant beta coefficient

**Table 8:** Multiple regression beta coefficients for MDC_95_% calculated using the following patient populations who have/are: Residential, Musculoskeletal, Hospitalised, and community dwellers. Vertically listed patient populations are the reference variables within the regression.

|  | Musculoskeletal | Hospitalised | Healthy<br>Community<br>dwellers |
| --- | --- | --- | --- |
| Residential | -5.784 | -4.927 | -10.728* |
| Musculoskeletal |  | 0.857 | -4.943 |
| Hospitalised |  |  | -5.800 |

\* denotes a statistically significant beta coefficient

## References

1. Baumgartner, R.N., Body composition in healthy aging. Annals of the New York Academy of Sciences, 2000. 904:437–48.

2. Pirker, W. and Katzenschlager, R., Gait disorders in adults and the elderly : A clinical guide. Wiener klinische Wochenschrift, 2017. 129:81–95.

3. Marcell, T.J., Sarcopenia: causes, consequences, and preventions. The journals of gerontology. Series A, Biological sciences and medical sciences, 2003. 58:M911–6.

4. Kojima, G., Iliffe, S., Jivraj, S. and Walters, K.J.J.E.C.H., Association between frailty and quality of life among community-dwelling older people: a systematic review and meta-analysis. Journal of Epidemiology and Community Health 2016. 70:716–721.

5. Heijink, A., et al., Biomechanical considerations in the pathogenesis of osteoarthritis of the knee. Knee Surgery, Sports Traumatology, Arthroscopy, 2012. 20:423–35.

6. Loeser, R.F., The role of aging in the development of osteoarthritis. Transactions of the American Clinical and Climatological Association, 2017. 128:44.

7. Malafarina, V., Uriz-Otano, F., Gil-Guerrero, L. and Iniesta, R., The anorexia of ageing: physiopathology, prevalence, associated comorbidity and mortality. A systematic review. Maturitas, 2013. 74:293–302.

8. World Health Organisation. Decade of healthy ageing: baseline report. 2021.

9. Auyeung, T.W., Lee, J.S., Kwok, T. and Woo, J., Physical frailty predicts future cognitive decline - a four-year prospective study in 2737 cognitively normal older adults. The Journal of Nutrition, Health & Aging, 2011. 15:690–4.

10. Middleton, A., et al., Self-Selected and Maximal Walking Speeds Provide Greater Insight Into Fall Status Than Walking Speed Reserve Among Community-Dwelling Older Adults. American Journal of Physical Medicine & Rehabilitation, 2016. 95:475–82.

11. Welmer, A.K., Rizzuto, D., Laukka, E.J., Johnell, K. and Fratiglioni, L., Cognitive and Physical Function in Relation to the Risk of Injurious Falls in Older Adults: A Population-Based Study. The Journals of Gerontology. Series A, Biological Sciences and Medical Sciences, 2017. 72:669–675.

12. Bohannon, R.W., et al., Comparison of walking performance over the first 2 minutes and the full 6 minutes of the Six-Minute Walk Test. BMC Research Notes, 2014. 7:269–269.

13. Bohannon, R.W., Bubela, D.J., Magasi, S.R., Wang, Y.-C. and Gershon, R.C., Sit-to-stand test: Performance and determinants across the age-span. Isokinetics and Exercise Science, 2010. 18:235–240.

14. Mancini, M. and Horak, F.B., The relevance of clinical balance assessment tools to differentiate balance deficits. European Journal of Physical and Rehabilitation Medicine, 2010. 46:239–48.

15. Reelick, M.F., van Iersel, M.B., Kessels, R.P. and Rikkert, M.G., The influence of fear of falling on gait and balance in older people. Age and Ageing, 2009. 38:435–40.

16. Ekstrom, H., Dahlin-Ivanoff, S. and Elmstahl, S., Effects of walking speed and results of timed get-up-and-go tests on quality of life and social participation in elderly individuals with a history of osteoporosis-related fractures. Journal of Aging and Health, 2011. 23:1379–99.

17. Sallinen, J., et al., Hand-grip strength cut points to screen older persons at risk for mobility limitation. Journal of theAmerican Geriatric Society, 2010. 58:1721–1726.

18. Fritz, S.L., Peters, D.M. and Greene, J.V., Measuring Walking Speed Clinical Feasibility and Reliability. Topics in Geriatric Rehabilitation, 2012. 28:91–96.

19. Rikli, R.E. and Jones, C.J., Functional fitness normative scores for community-residing older adults, ages 60-94. Journal of Aging and Physical Activity, 1999. 7:162–181.

20. Hopkins, W.G., Measures of Reliability in Sports Medicine and Science. Sports Medicine, 2000. 30:1–15.

21. Haley, S.M. and Fragala-Pinkham, M.A.J.P.t., Interpreting Change Scores of Tests and Measures used in Physical Therapy. Physical Therapy, 2006. 86:735–743.

22. Chui, K., Hood, E. and Klima, D., Meaningful Change in Walking Speed. Topics in Geriatric Rehabilitation, 2012. 28:97–103.

23. Hornyak, V., VanSwearingen, J.M. and Brach, J.S., Measurement of Gait Speed. Topics in Geriatric Rehabilitation, 2012. 28:27–32.

24. Rydwik, E., Bergland, A., Forsen, L. and Frandin, K., Investigation into the Reliability and Validity of the Measurement of Elderly People’s Clinical Walking Speed: a Systematic Review. Physiotherapy Theory and Practice, 2012. 28:238–56.

25. Downs, S., Marquez, J. and Chiarelli, P., The Berg Balance Scale has High Intra-and Inter-rater Reliability but Absolute Reliability Varies Across the Scale: a Systematic Review. Journal of Physiotherapy, 2013. 59:93–9.

26. Pin, T.W., Psychometric Properties of 2-Minute Walk Test: A Systematic Review. Archives of Physical Medicine and Rehabilitation, 2014. 95:1759–1775.

27. Bohannon, R.W., Minimal Clinically Important Difference for Grip Strength: a Systematic Review. Journal Physical Therapy Science, 2019. 31:75–78.

28. Bohannon, R.W. and Crouch, R., Minimal Clinically Important Difference for Change in 6-minute Walk Test Distance of Adults with Pathology: a Systematic Review. Journal of Evaluation in Clinical Practice, 2017. 23:377–381.

29. Kotter-Gruhn, D., Neupert, S.D. and Stephan, Y., Feeling Old Today? Daily Health, Stressors, and Affect Explain Day-to-Day Variability in Subjective Age. Psychological Health, 2015. 30:1470–85.

30. Weir, J., Quantifying Test-Retest Reliability Using The Intraclass Correlation Coefficient and the SEM. Journal of Strength and Conditioning Research, 2005. 19:231–40.

31. Choi, Y.M., Dobson, F., Martin, J., Bennell, K.L. and Hinman, R.S., Interrater and Intrarater Reliability of Common Clinical Standing Balance Tests for People With Hip Osteoarthritis. Physical Therapy, 2014. 94:696–704.

32. Konig, N., Taylor, W.R., Baumann, C.R., Wenderoth, N. and Singh, N.B., Revealing the Quality of Movement: A Meta-Analysis Review to Quantify the Thresholds to Pathological Variability During Standing and walking. Neuroscience & Biobehavioral Reviews 2016. 68:111–119.

33. Hansen, H., Beyer, N., Frolich, A., Godtfredsen, N. and Bieler, T., Intra-and Inter-Rater Rof the 6-minute WTest and the 30-second Sit-to-Stand test in Patients with Severe and Very Severe COPD. International Journal of Chronic Obstructive Pulmonary Disease, 2018. 13:3447–3457.

34. Labadessa, I.G., et al., Should the 6-Minute Walk Test Be Compared When Conducted by 2 Different Assessors in Subjects With COPD? Respiratory Care, 2016. 61:1323–30.

35. Dodds, R.M., et al., Grip Strength Across the Life Course: Normative Data from Twelve British Studies. PloS One, 2014. 9:e113637.

36. Mokkink, L.B., et al., COSMIN Risk of Bias tool to Assess the Quality of Studies on Reliability or Measurement Error of Outcome Measurement Instruments: a DelphiStudy. BMC Medical Research Methodology, 2020. 20:293.

37. Guralnik, J., et al., Clinically Meaningful Change for Physical Performance: Perspectives of the ICFSR Task Force. The Journal of Frailty & Aging, 2020. 9: 9–13.

38. Barrios-Fernandez, S., et al., Reliability of 30-s Chair Stand Test with and without Cognitive Task in People with Type-2 Diabetes Mellitus. International Journal of Environmental Research and Public Health, 2020. 17.

39. Bodilsen, A.C., et al., Feasibility and Inter-Rater Reliability of Physical Performance Measures in Acutely Admitted Older Medical Patients. PloS One, 2015. 10.

40. Unver, B., et al., Test-retest Reliability of the 50-foot Timed Walk and 30-second Chair Stand Test in Patients with Total Hip Arthroplasty. Acta Orthopaedica Belgica, 2015. 81:435–41.

41. Dobson, F., et al., Reliability and Measurement Error of the Osteoarthritis Research Society International (OARSI) Recommended Performance-based Tests of Physical Function in People with Hip and Knee Osteoarthritis. Osteoarthritis and Cartilage, 2017. 25:1792–1796.

42. Bohannon, R.W., The PhyStat 7: A New Test Battery for Characterizing the Physical Status of Older Adults. Topics in Geriatric Rehabilitation, 2017. 33:84–88.

43. Goldberg, A., Chavis, M., Watkins, J. and Wilson, T., The Five-times-sit-to-stand Test: Falidity, Reliability and Detectable Change in Older Females. Aging Clinical and Experimental Research, 2012. 24:339–344.

44. Altubasi, I.M., Determine Real Change in Performance During Single session of Functional Testing. Journal of Musculoskeletal & Neuronal Interactions, 2019. 19:215–219.

45. Melo, T.A., et al., The Five Times Sit-to-Stand Test: Safety and Reliability with Older Intensive Care Unit Patients at Discharge. Revista Brasileira de terapia intensiva, 2019. 31:27–33.

46. Silva, A.G., et al., Inter-rater Reliability, Standard Error of Measurement and Minimal Detectable Change of the 12-item WHODAS 2.0 and Four Performance Tests in Institutionalized Ambulatory Older Adults. Disability and Rehabilitation, 2017:1–8.

47. Doll, H., Gentile, B., Bush, E.N. and Ballinger, R., Evaluation of the Measurement Properties of Four Performance Outcome Measures in Patients with Elective Hip Replacements, Elective Knee Replacements, or Hip Fractures. Value in health : the Journal of the International Society for Pharmacoeconomics and Outcomes Research, 2018. 21:1104–1114.

48. Medina-Mirapeix, F., et al., Five Times Sit-to-Stand Test in Subjects with Total Knee Replacement: Reliability and Relationship with Functional Mobility Tests. Gait & Posture, 2018. 59:258–260.

49. Lin, Y.C., Davey, R.C. and Cochrane, T., Tests for Physical Function of the Elderly with Knee and Hip Osteoarthritis. Scandinavian Journal of Medicine & Science in Sports, 2001. 11:280–286.

50. Mangione, K.K., et al., Detectable Changes in Physical Performance Measures in Elderly African Americans. Physical Therapy, 2010. 90:921–927.

51. Mathis, R.A., Taylor, J.D., Odom, B.H. and Lairamore, C., Reliability and Validity of the Patient-Specific Functional Scale in Community-Dwelling Older Adults. Journal of Geriatric Physical Therapy (2001), 2019.42:E67–E72

52. Lee, S.P., Dufek, J., Hickman, R. and Schuerman, S., Influence of Procedural Factors on the Reliability and Performance of the Timed Up-and-go Test in Older Adults. International Journal of Gerontology, 2016. 10:37–42.

53. Goldberg, A. and Schepens, S., Measurement Error and Minimal Detectable Change in Timed Up and Go in Older Adults. Gerontologist, 2010. 50:337–337.

54. Kristensen, M.T., Bloch, M.L., Jonsson, L.R. and Jakobsen, T.L., Interrater Reliability of the Standardized Timed Up and Go Test when used in Hospitalized and Community-dwelling Older Individuals. Physiotherapy Research International, 2019. 24.

55. Suzuki, Y., et al., Absolute Reliability of Measurements of Muscle Strength and Physical Performance Measures in Older People with High Functional Capacities. European Geriatric Medicine, 2019. 10:733–740.

56. Kristensen, M.T., Henriksen, S., Stie, S.B. and Bandholm, T., Relative and Absolute Intertester Reliability of the Timed Up and Go Test to Quantify Functional Mobility in Patients with Hip Fracture. Journal of the American Geriatrics Society, 2011. 59:565–7.

57. Naylor, J.M., et al., Minimal Detectable Change for Mobility and Patient-reported Tools in People with Osteoarthritis Awaiting Arthroplasty. BMC Musculoskeletal Disorders, 2014. 15.

58. Yeung, T.S., Wessel, J., Stratford, P.W. and MacDermid, J.C., The Timed Up and Go Test for Use on an Inpatient Orthopaedic Rehabilitation Ward. The Journal of Orthopaedic and Sports Physical Therapy, 2008. 38:410–7.

59. Yuksel, E., Kalkan, S., Cekmece, S., Unver, B. and Karatosun, V., Assessing Minimal Detectable Changes and Test-Retest Reliability of the Timed Up and Go Test and the 2-Minute Walk Test in Patients With Total Knee Arthroplasty. Journal of Arthroplasty, 2017. 32:426–430.

60. Kennedy, D.M., Stratford, P.W., Wessel, J., Gollish, J.D. and Penney, D., Assessing Stability and Change of Four Performance Measures: a Longitudinal Study Evaluating Outcome Following Total Hip and Knee Arthroplasty. BMC Musculoskeletal Disorders, 2005. 6.

61. Faleide, A.G.H., Bogen, B.E. and Magnussen, L.H., Intra-session Test-retest Reliability of the Timed “Up & Go” Test when Performed by Patients with Hip Fractures. European Journal of Physiotherapy, 2015. 17:89–97.

62. Goldberg, A., Casby, A. and Wasielewski, M., Minimum Detectable Change for Single-leg-stance-time In older adults. Gait & Posture, 2011. 33:737–739.

63. Ziv, E., Patish, H. and Dvir, Z., Grip and Pinch Strength in Healthy Subjects and Patients with Primary Osteoarthritis of the Hand: a Reproducibility Study. Open Orthopaedics Journal, 2008. 2:86–90.

64. Jenkins, N.D., et al., Reliability and Jelationships among Handgrip Strength, Leg Extensor Strength and Power, and Balance in Older Men. Experimental Gerontology, 2014. 58:47–50.

65. Jenkins, N.D.M. and Cramer, J.T., Reliability and Minimum Detectable Change for Common Clinical Physical Function Tests in Sarcopenic Men and Women. Journal of the American Geriatriac Society, 2017. 65:839–846.

66. Telenius, E.W., Engedal, K. and Bergland, A., Inter-rater Reliability of the Berg Balance Scale, 30 s Chair Stand Test and 6 m Walking Test, and Construct Validity of the Berg Balance Scale in Nursing Home Residents with Mild-to-Moderate Dementia. BMJ Open, 2015. 5.

67. Goldberg, A. and Schepens, S., Measurement Error and Minimum Detectable Change in 4-meter Gait Speed in Older Adults. Aging Clinical and Experimental Research, 2011. 23:406–412.

68. Forte, R., De Vito, G. and Boreham, C.A.G., Reliability of Walking Speed in Basic and Complex Conditions in Healthy, Older Community-dwelling Individuals. Aging Clinical and Expimental Research, 2020.

69. Hollman, J.H., et al., Minimum Detectable Change in Gait Velocity During Acute Rehabilitation Following Hip Fracture. Journal of Geriatric Physical Therapy (2001), 2008. 31:53–6.

70. Barthuly, A.M., Bohannon, R.W. and Gorack, W., Gait Speed is a Responsive Measure of Physical Performance for Patients Undergoing Short-term Rehabilitation. Gait & Posture, 2012. 36:61–64.

71. Fransen, M., Crosbie, J. and Edmonds, J., Reliability of Gait Measurements in People with Osteoarthritis of the Knee. Physical Therapy, 1997. 77:944–53.

72. Fernández-Huerta, L. and Córdova-León, K., Reliability of Two Gait Speed Tests of Different Timed Phases and Equal Non-timed Phases in Community-dwelling Older Persons. Medwave, 2019. 19:e7611.

73. Overgaard, J.A., Larsen, C.M., Holtze, S., Ockholm, K. and Kristensen, M.T., Interrater Reliability of the 6-Minute Walk Test in Women With Hip Fracture. Journal of Geriatric Physical Therapy, 2017. 40:158–166.

74. Chan, W.L.S. and Pin, T.W., Reliability, Validity and Minimal Detectable Change of 2-min Walk Test and 10-m Walk Test in Frail Older Adults Receiving Day Care and Residential Care. Aging Clinical and Experimental Research, 2020. 32:597–604.

75. Connelly, D.M., Thomas, B.K., Cliffe, S.J., Perry, W.M. and Smith, R.E., Clinical Utility of the 2-minute Walk Test for Older Adults Living in Long-term Care. Physiotherapy Canada, 2009. 61:78–87.

